# Proximal and distal factors predicting timely initiation of breast feeding in Ethiopia: a systematic review and meta-analysis

**DOI:** 10.1101/19000497

**Authors:** Tesfa Dejenie Habtewold, Shimels Hussien Mohammed, Aklilu Endalamaw, Henok Mulugeta, Getenet Dessie, Derbew Fikadu Berhe, Mulugeta Molla Birhanu, Md. Atiqul Islam, Andreas A. Teferra, Nigus Gebremedhin Asefa, Sisay Mulugeta Alemu

## Abstract

**Background:** In Ethiopia, the current coverage of timely initiation of breast feeding (TIBF) has fallen short of the national Health Sector Transformation Plan 2016-2020, National Nutrition Program 2016–2020 and WHO global target. This calls for the need to assess relevant proximal and distal factors that affect the rate of TIBF in Ethiopia.

**Objective:** The aim of this meta-analysis was to investigate the association between TIBF and educational status, household income, marital status, media exposure, and parity in Ethiopia.

**Methods:** Databases used were PubMed, EMBASE, Web of Science, SCOPUS, CINAHL and WHO Global health library, and key terms were searched using interactive searching syntax. It was also supplemented by manual searching. Observational studies published between September 2000 and March 2019 were included. The methodological quality of studies was examined using the Newcastle-Ottawa Scale (NOS) for cross-sectional studies. Data were extracted using the Joanna Briggs Institute (JBI) data extraction tool. To obtain the pooled odds ratio (OR), extracted data were fitted in a random-effects meta-analysis model. Statistical heterogeneity was quantified using Cochran’s Q test, τ^2^, and I^2^ statistics. Additional analysis conducted includes Jackknife sensitivity analysis, cumulative meta-analysis, and meta-regression analysis.

**Results:** Out of 553 studies retrieved, 25 studies fulfilled our inclusion criteria. Almost all studies were conducted on mothers with newborn less than 23 months. Maternal educational status (OR = 1.82; p < 0.001; 95% CI = 1.35 - 2.45; I^2^ = 84.96%), paternal educational status (OR = 2.72; p = 0.001, 95% CI = 1.49 - 4.97 I^2^ = 62.50%), income (OR = 1.16; p = 0.002; 95% CI = 1.05 - 1.27; I^2^ = 0.00%), marital status (OR = 1.39; p = 0.001; 95% CI = 1.14 - 1.69; I^2^ = 9.17%) and parity (OR = 1.39; p = 0.01; 95% CI = 1.07 - 1.81; I^2^ = 74.43%) were found to be significantly associated with TIBF. We also observed a direct dose-response relationship of TIBF with educational status and income.

**Conclusions:** Proximal and distal factors significantly predicting TIBF practice in Ethiopia, which needs integrated intervention by health professionals and healthcare policymakers. Health education, counselling and peer education targeting parents at antenatal and postnatal periods are needed. It is also relevant to improve the economic power of women and promote gender equality.

## Background

Timely initiation of breast feeding (TIBF) is defined as putting the newborn to the breast within the first-hour birth.(1, 2) Based on the recent UNICEF and World Health Organization (WHO) global report, approximately 78 million newborns are not breastfeed within the first hour after birth; majority are born in low- and middle-income countries (LMICs).(3) Breast feeding with timely initiation and optimal duration is a core component of primary healthcare that boosts newborn later life health (e.g. cognitive function)(4), reduce infection, and prevent up to 60% infant and child morbidity and mortality related to malnutrition.(5-11) Recent estimates suggest that optimal breast feeding could prevent approximately 12% deaths per year in under-five children, which represents around 800,000 lives in LMICs.(12) Likewise, TIBF has been shown to reduce neonatal mortality by 22%.(7) Breast feeding also prevents acute and chronic maternal diseases.(13, 14) Optimal breast feeding has also economic and environmental advantages.(13)

Suboptimal breast feeding including delayed initiation increases risk of newborn morbidity and mortality up to fivefold.(5, 15-17) The 2010 Global Burden of Disease study reveals that suboptimal breast feeding is one of the three leading causes of disease in Sub-Saharan African (SSA) countries.(18) Approximately 25 to 50% of infant deaths in developing countries occur due to poor breast feeding practice.(16, 19) In terms of the global absolute number of under-five child deaths, Ethiopia ranked sixth with about 472,000 deaths per year. The age distribution of under-five deaths varies from 29% in the first 30 days of life to 42% in the fourth year.(20) Neonatal mortality rate is still high in Ethiopia, which accounts for 42% of under-five deaths.(21, 22) Based on our previous meta-analysis of 13 observational studies conducted in Ethiopia, the prevalence of neonatal mortality is 16.3%.(23)

To date, only 22 countries achieved WHO target of 70% coverage in TIBF.(24) Although LMICs have strong breast feeding traditions compared to developed countries, adherence to WHO recommendations of TIBF is poor with considerable difference between nations. A report from 76 countries on the rates and trends of TIBF over the past decade shows that TIBF rates varied widely across regions from 32% in East Asia and the Pacific to 65% in Eastern and Southern Africa.(25) A demographic and health survey conducted in 57 LMICs involving nearly 200,000 newborn shows that 39% of newborn (with regional range from 31 to 60%) breastfed within one hour of birth.(26) Likewise, a report of demographic and health surveys conducted since 2010 in 32 SSA countries show that the prevalence of TIBF is 50.5%.(27) A systematic review of studies conducted between 1999 and 2013 in Asia, Africa, and South America reveal the prevalence of TIBF ranges from 11.4% in a province of Saudi Arabia to 83.3% in Sri Lanka.(28) In Ethiopia, breast feeding is a common practice with 96% of mothers breastfeed at least once during their lifetime.(29) According to our previous meta-analysis of 45 observational studies, the prevalence of TIBF in Ethiopia is 66.5%.(30)

Numerous country-specific or global factors influence mothers’ decision and ability to initiate breast feeding within one hour of birth. The accumulated body of evidence from regional and global studies consistently shows that TIBF has been associated with numerous factors including educational status, household income, marital status, media exposure and parity.(31-39) In our previous systematic review of 70 observational studies, we identified 18 predictors categorized into four groups: proximal (maternal occupational status, maternal knowledge on TIBF, and breast feeding guidance and counselling), proximal-intermediate (place of delivery, mode of delivery, birth attendant and sex of newborn), distal-intermediate (antenatal care, postnatal care, prelacteal feeding, and colostrum feeding) and distal (paternal educational status, household income, marital status, media exposure, family size, breast feeding experience, place of residence, birth order, parity, and iron-folate supplementation).(30) In the follow-up meta-analyses(30, 40, 41), we confirmed that breast feeding guidance and counselling, mode of delivery, place of delivery, antenatal care visit and colostrum discarding significantly associated with TIBF practice, whereas maternal occupational status, maternal or caregiver’s age and sex of newborn not significantly associated with TIBF.

Given the multidimensional benefits of breast feeding for the newborn, mother and community, global initiatives have been made to improve the national and international breast feeding practice. Some of these initiatives are the International Code of Marketing of Breast-milk Substitutes (aka the Code), Innocenti Declaration, Baby Friendly Hospital Initiative (BFHI) (i.e. Ten Steps to Successful Breast feeding), Millennium Development Goals, Global Nutrition Targets 2025 and Sustainable Development Goals.(31, 42) The Ethiopian government has endorsed and implemented these aforementioned policies and programs to reduce infant and child mortality and morbidity related to poor breast feeding in the country. Since 2004, the Ethiopian Federal Ministry of Health has developed infant and young child feeding guidelines and provided breast feeding promotions.(43) A national nutrition strategy and program (NNP) has also been developed and implemented in a multi-sectoral approach. To improve the nutritional status of mothers and children, the Health Sector Development Plan (HSDP)-IV has integrated nutrition into the health extension program.(44)

Consequently, the rate of malnutrition, stunting, underweight and under-five mortality have declined in Ethiopia.(45) Despite the heartening reduction of under-five morbidity and mortality, the rate of TIBF has fallen short of the national Health Sector Transformation Plan 2016-2020(44), National Nutrition Program 2016–2020(46) and WHO global target.(24) This calls for the need to investigate the pooled effect of various proximal and distal factors predicting TIBF. Therefore, the aim of this meta-analysis was to investigate the association between TIBF and maternal educational status, paternal educational status, household income, marital status, media exposure, and parity in Ethiopia. We hypothesized high educational status, high household income, being married, exposed to media and multiparity significantly increase the odds of TIBF.

## Methods

### Measurements

The outcome measurement was timely initiation of breast feeding (TIBF), which is the proportion of children born in the last two years who were breastfeed within the first hour of delivery.(47) Maternal educational status, paternal educational status, household income, marital status, media exposure, and parity were selected predicting factors. The selection of these factors was guided by our previous systematic review findings.(30) Educational status represents the highest schooling grade achieved as per the Ethiopian educational system and categorized as ‘uneducated’ (including these who able to read and write without formal schooling), ‘primary’ (grades 1 to 8) and ‘secondary and above’ (grades 9 or above). Household income was categorized as ‘high’, ‘medium’ and ‘low’. Because of substantial inconsistency in the reported household income, we used a harmonized qualitative way of classification of income for all included studies that reported at least three categories of household income or wealth index based on authors educational judgment. Marital status was categorized as ‘currently married’ and ‘others’ (i.e. single, divorced, widowed). Media exposure represents exposure to or ownership of any print media (newspaper, leaflet, brochure) and broadcasting (radio and television) and categorized as ‘yes’ and ‘no’. This does not include Facebook, email, YouTube, or WhatsApp as most mothers do not have access to internet. If studies reported accessibility or exposure to more than one media tools, we extracted data on radio (as ‘yes’) followed by television and print media. Parity refers to the total number of births after 28 weeks and was categorized as ‘primipara’ if the mothers have only one birth and ‘multipara’ if the mothers have at least two births.

### Search for literature

Databases searched were PubMed, EMBASE, Web of Science, SCOPUS, CINAHL and WHO Global health library. The interactive searching syntax was developed for all databases in consultation with librarian, experts on literature searching (Supplementary file 1). We further manually searched the table of contents of Ethiopian Journal of Health Development, Ethiopian Journal of Health Sciences, Ethiopian Journal of Reproductive Health, International Breast feeding Journal, BMC Pregnancy and Childbirth, BMC Public Health, BMC Paediatrics, Nutrition Journal and Italian Journal of Paediatrics. We also searched cross-references and grey literature on Addis Ababa University institutional research collection repository database. The last search was done in March 2019.

### Inclusion and exclusion criteria

The studies were included when they met the following inclusion criteria: (1) observational studies, such as cross-sectional, case-control and cohort studies; (2) studies conducted in Ethiopia; (3) studies reported on the association between TIBF (i.e. operationalized based on the WHO definition) and maternal and paternal educational status, household income (at least three categories must be reported), marital status, media exposure (not exposed or no access to media category must be reported) and parity; (4) studies published from September 2000 (i.e. the time when last revision of the global breast feeding recommendations occurred) to March 2019. Program evaluation reports, systematic reviews and meta-analyses, qualitative studies and studies on mothers with medical conditions including HIV/AIDS and pre-term or ill health newborn were excluded.

### Screening and selection of studies

Initially, all identified studies were exported into RefWorks citation manager version 4.6 for Windows. Afterward, duplicate studies were deleted from further screening. Next, a pair of reviewers independently screened the abstracts and titles using Microsoft Excel spreadsheet for relevance, their compliance with our measurements of interest and against our inclusion criteria. Based on Cohen’s Kappa inter-rater reliability test, the agreement between the two reviewers was 0.76 indicating substantial agreement between the two reviewers. Disagreements on the inclusion of titles or abstracts were solved through discussion and consensus. After removing irrelevant studies, full text of selected abstracts were downloaded and reviewed for further eligibility. PRISMA flow diagram was also used to illustrate the screening and selection processes of studies.(48) Finally, two independent reviewers (TD and SM) extracted the following information from each included studies using Joanna Briggs Institute (JBI) data extraction tool(49): author name, publication year, residence, study design, study population, number of participants, source of funding, and observed data. If funding source was not explicitly mentioned, we reported as ‘not mentioned’, whereas ‘no funding’ category was used if the author explicitly mentioned there is no funding or funding not applicable. If funding was not given directly but through other donors, the original donor was mentioned. Newcastle-Ottawa Scale (NOS) for cross-sectional studies was used to examine the quality of studies and the potential risk of bias.(50) The scoring system, selection of cut-off value and interpretation used in this meta-analysis were published in our protocol.(51) Furthermore, we reported the results in compliance with the recommendation of Preferred Reporting Items for Systematic review and Meta-analysis (PRISMA) statement (Supplementary file 2).(48)

### Statistical analysis

To obtain the pooled odds ratio (OR), extracted data were fitted in a random-effects meta-analysis model. In addition, a cumulative meta-analysis was done to illustrate the trend of evidence regarding the effect of predictors on TIBF and interpreted as stable, steadily increased/decreased, slightly increased/decreased or dramatically increased/decreased. Publication bias was assessed by subjective evaluation of the funnel plot, and then, we performed Egger’s regression statistical test to objectively confirm the presence of significant publication bias at p-value ≤ 0.01.(52) We used Cochran’s Q test to test heterogeneity, τ^2^ to estimate the amount of total/residual between-study variance, and I^2^ statistics to measure the proportion of total variation between study due to heterogeneity.(53) Clinical and methodology heterogeneity were also carefully evaluated. Factors attributed to between-study heterogeneity were investigated using mixed-effects meta-regression analysis using region, residence, sample size and publication year as covariates.(54) The residual amount of heterogeneity was subtracted from the proportion of heterogeneity and divided by the total amount of heterogeneity to obtain the total amount of heterogeneity (R^2^) explained by covariates. Omnibus test of moderators was applied to assess the moderation effect of these covariates. Meta-regression analysis was done only when heterogeneity threshold (I^2^) was ≥ 80%. Jackknife sensitivity analysis was done to examine the influence of outlier studies on the pooled OR estimate, a significance level of estimate and between-study heterogeneity.(55) The study was excluded when the pooled OR estimate increased or decreased by one and changes the significance level after lifting out that study from the meta-analysis. Because of the small number of studies available for some variables, the change in heterogeneity threshold was not considered as a primary criterion to detect and excluded the outlier study. The data were analyzed using “metafor” packages in R software version 3.2.1 for Windows.(54)

### Review protocol

This systematic review and meta-analysis was conducted based on the registered (CRD42017056768) and published protocol.(51) Based on the authors’ decision, the following changes were made to the published protocol.(51) Joanna Briggs Institute (JBI) tool(49) was used to extract the relevant data. Furthermore, cumulative meta-analysis and mixed-effects meta-regression analysis were done to reveal the trends of evidence and identify possible sources of between-study heterogeneity respectively.

## Results

### Search results

In total, 483 studies through electronic databases and 70 studies through manual searching were retrieved for further screening. Following a rigorous screening, 28 studies were selected for full text review. Three studies were excluded after the full text review: one study(56) reported only the prevalence of TIBF, and the other two studies(20, 57) did not report the selected factors of our interest. A total of 25 studies were included in this meta-analysis, which most of them conducted on mothers with newborn less than 23 months. All studies had good methodological quality (NOS score ≥7). One study reported more than one predicting factors. The screening and selection process are illustrated below using the PRISMA flow diagram (Figure 1). The detailed characteristics of studies that reported each variable is presented in Table 1.

**Figure 1.**
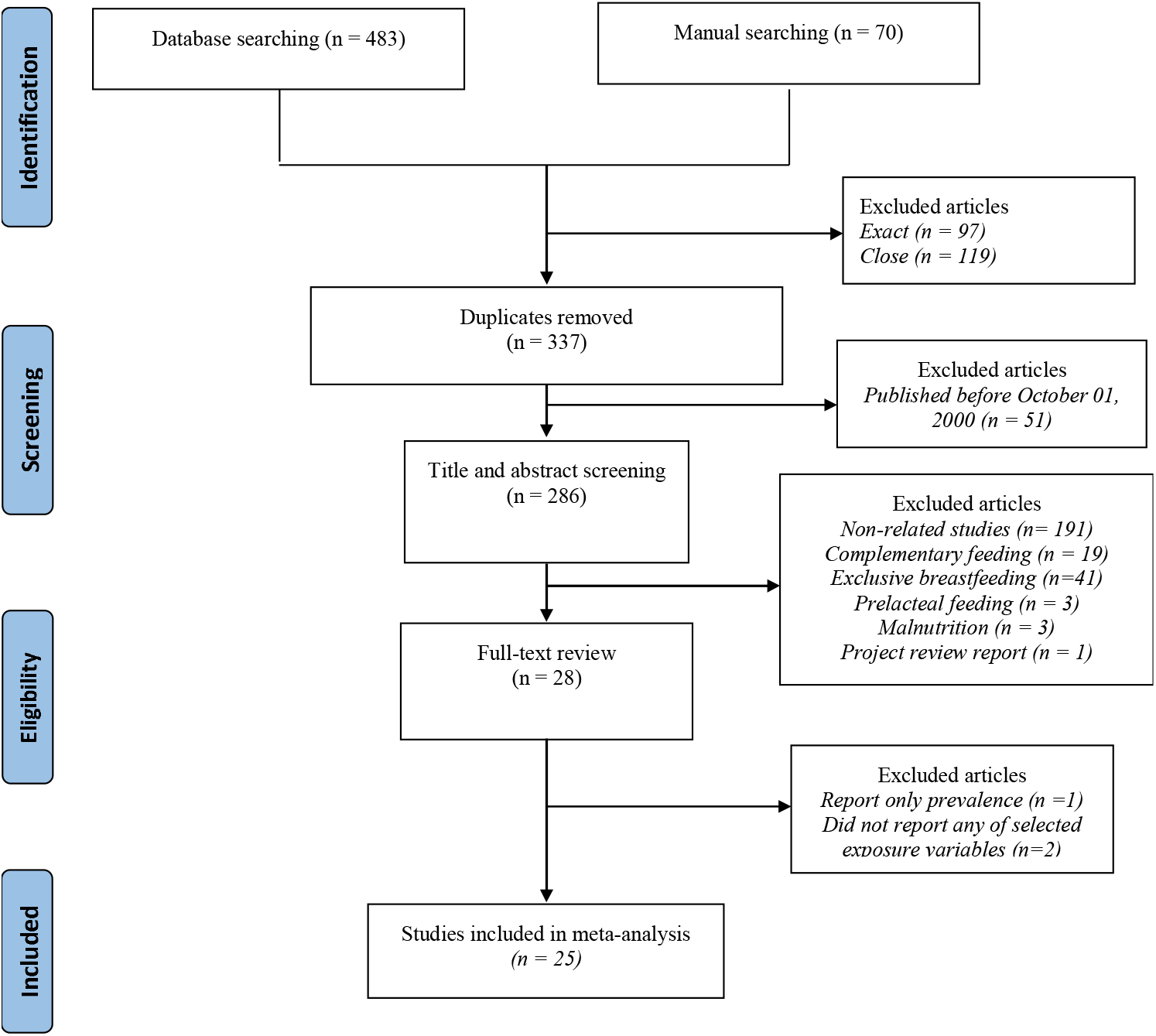
PRISMA flow diagram showing the schematics of literature screening and selection.

**Table 1:**
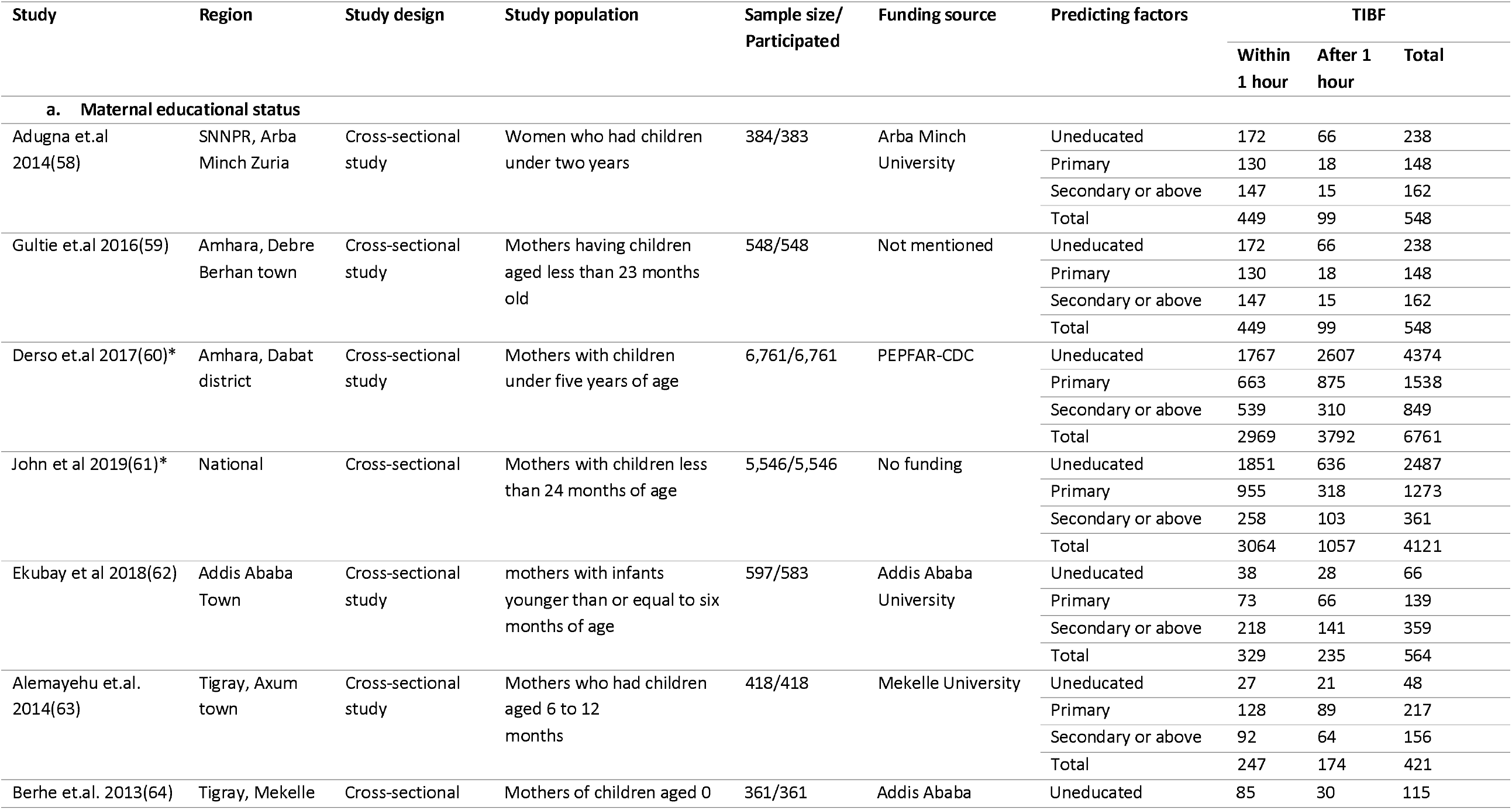
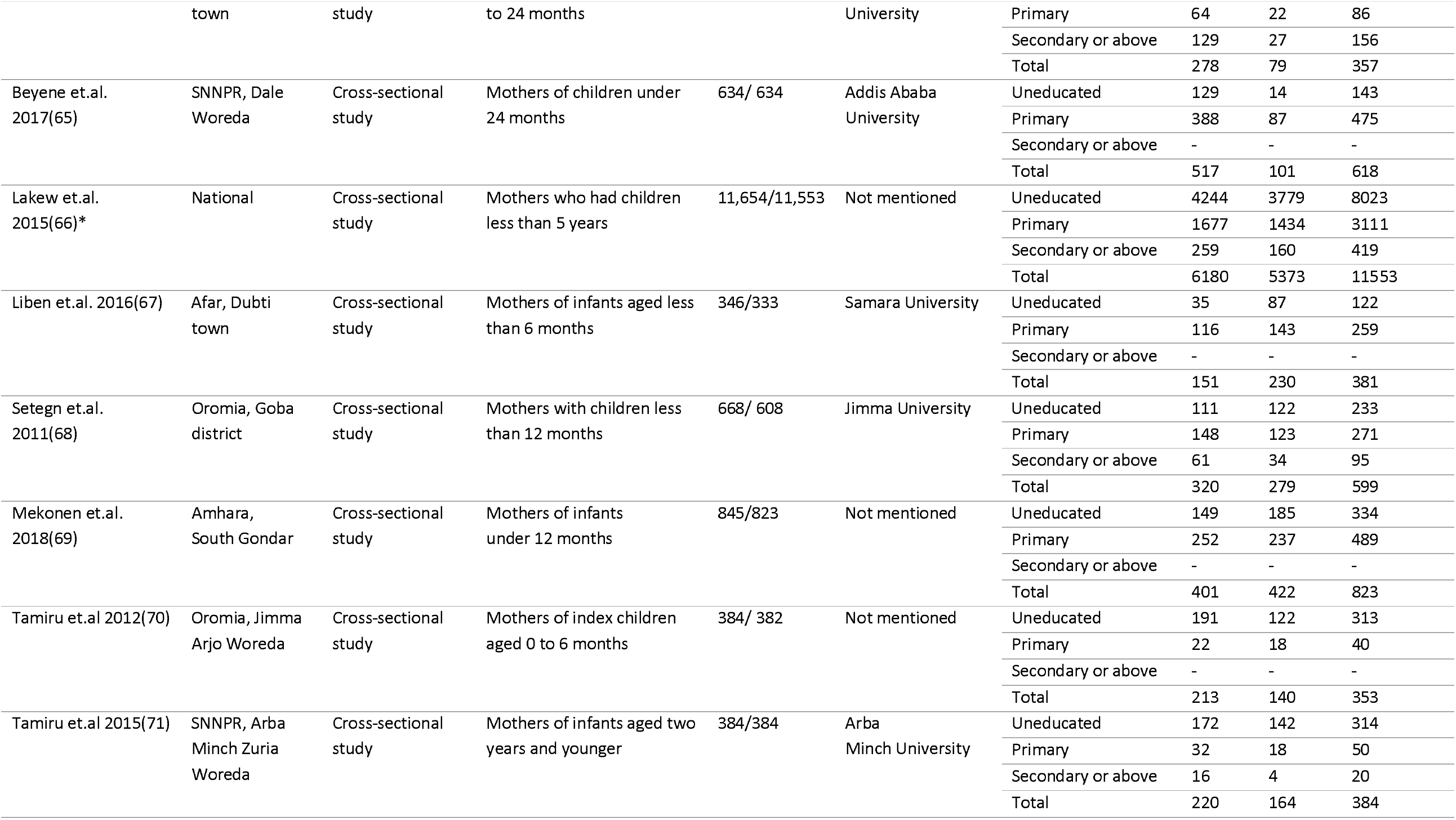
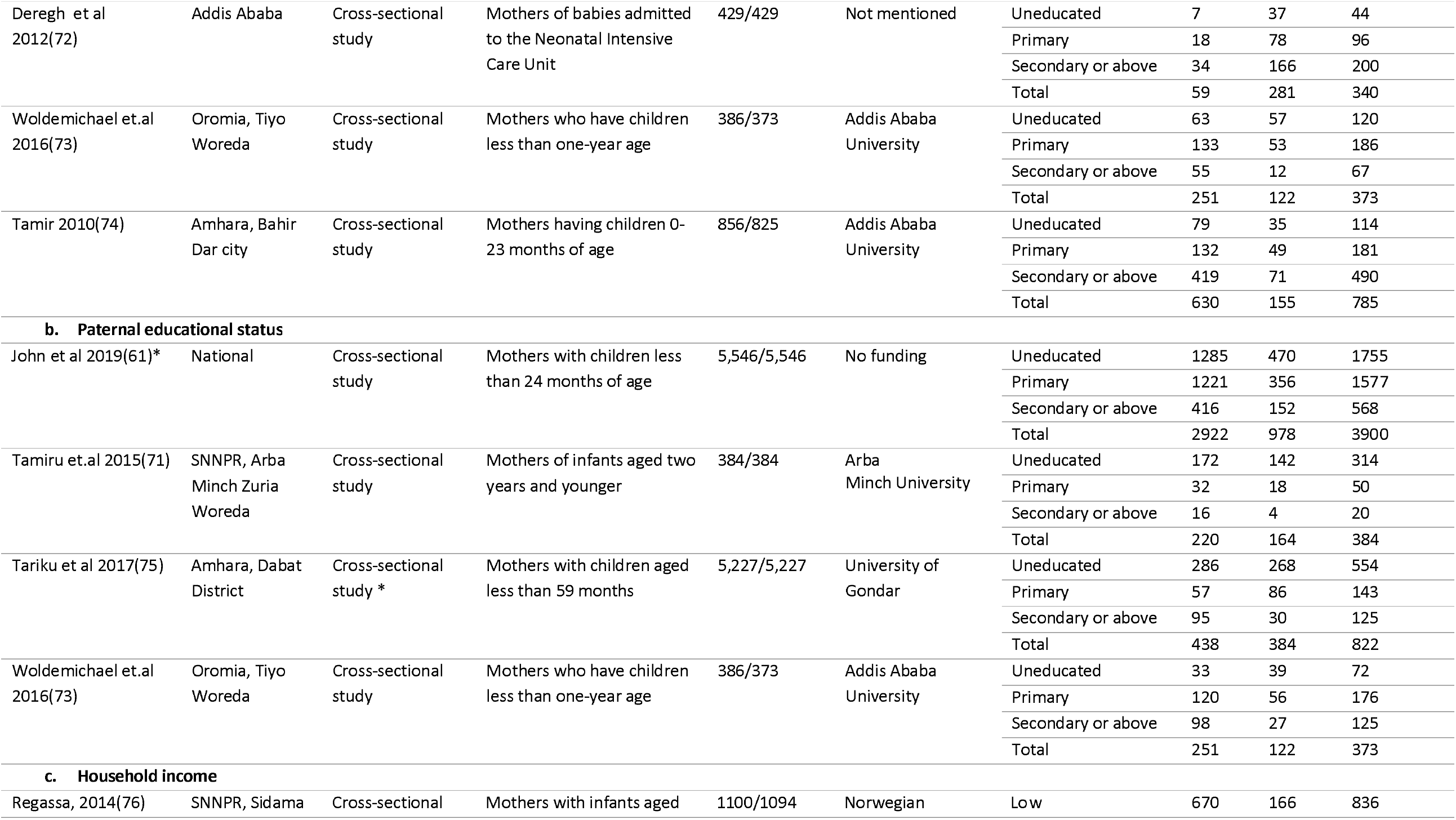
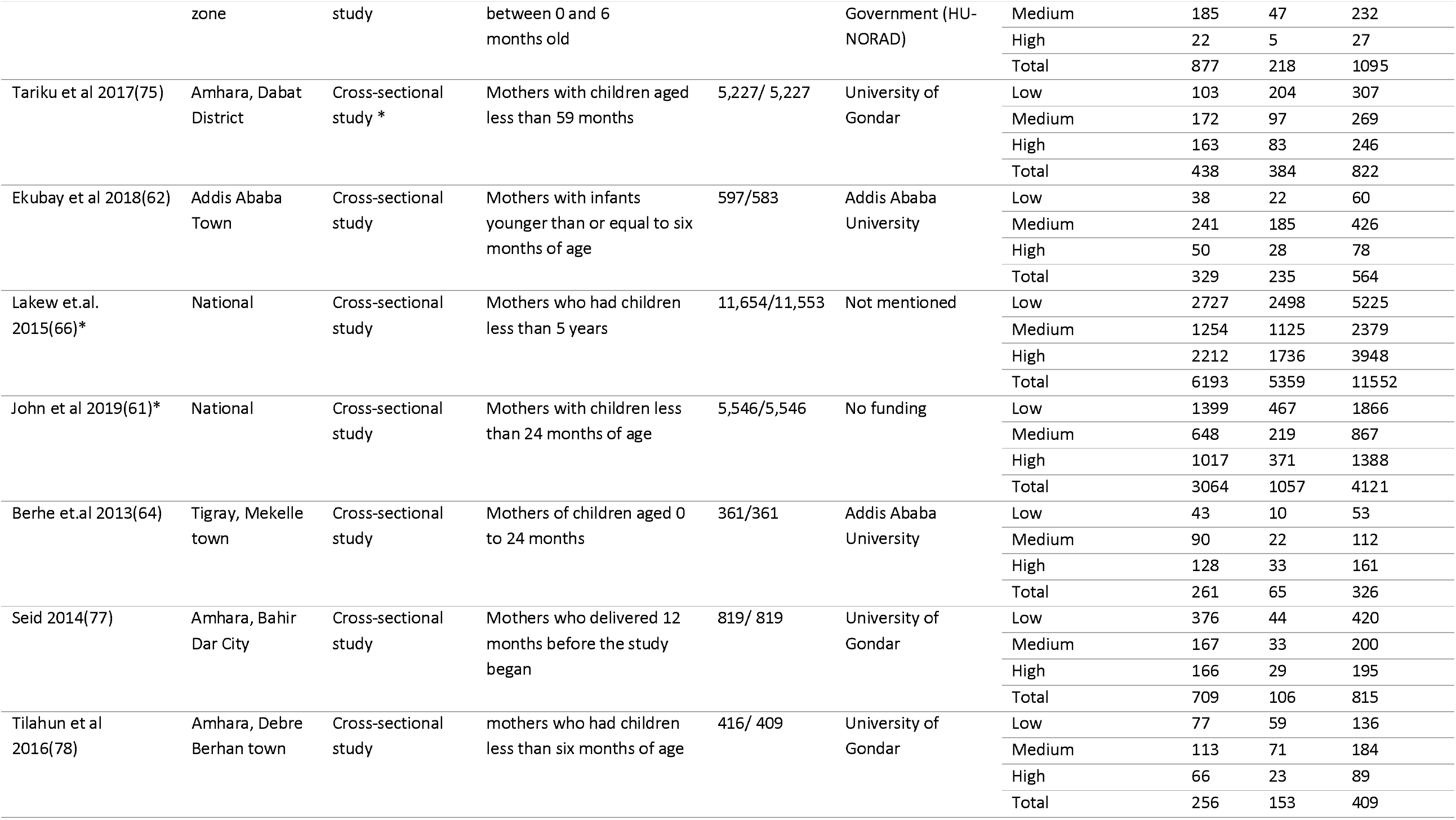
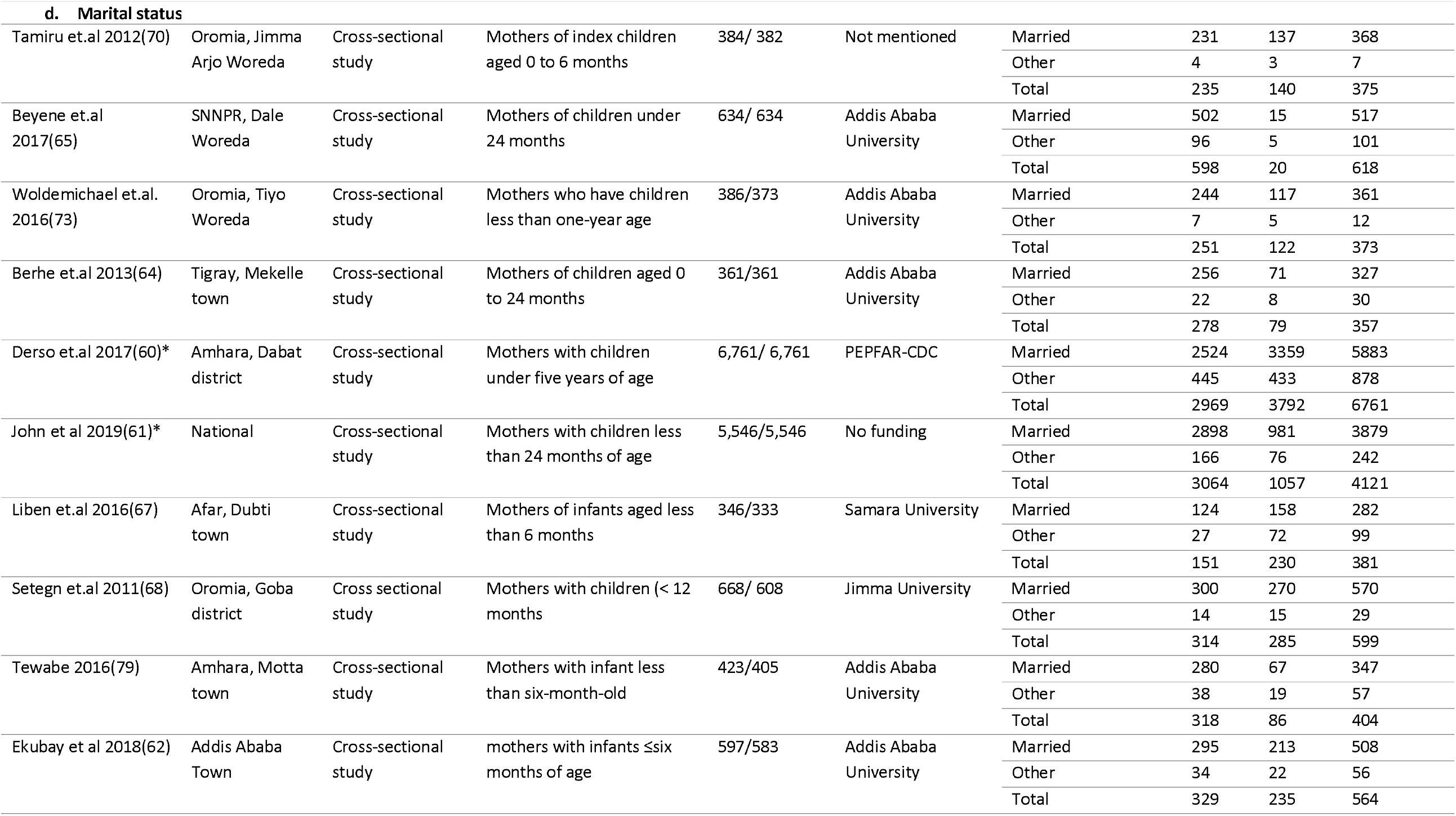
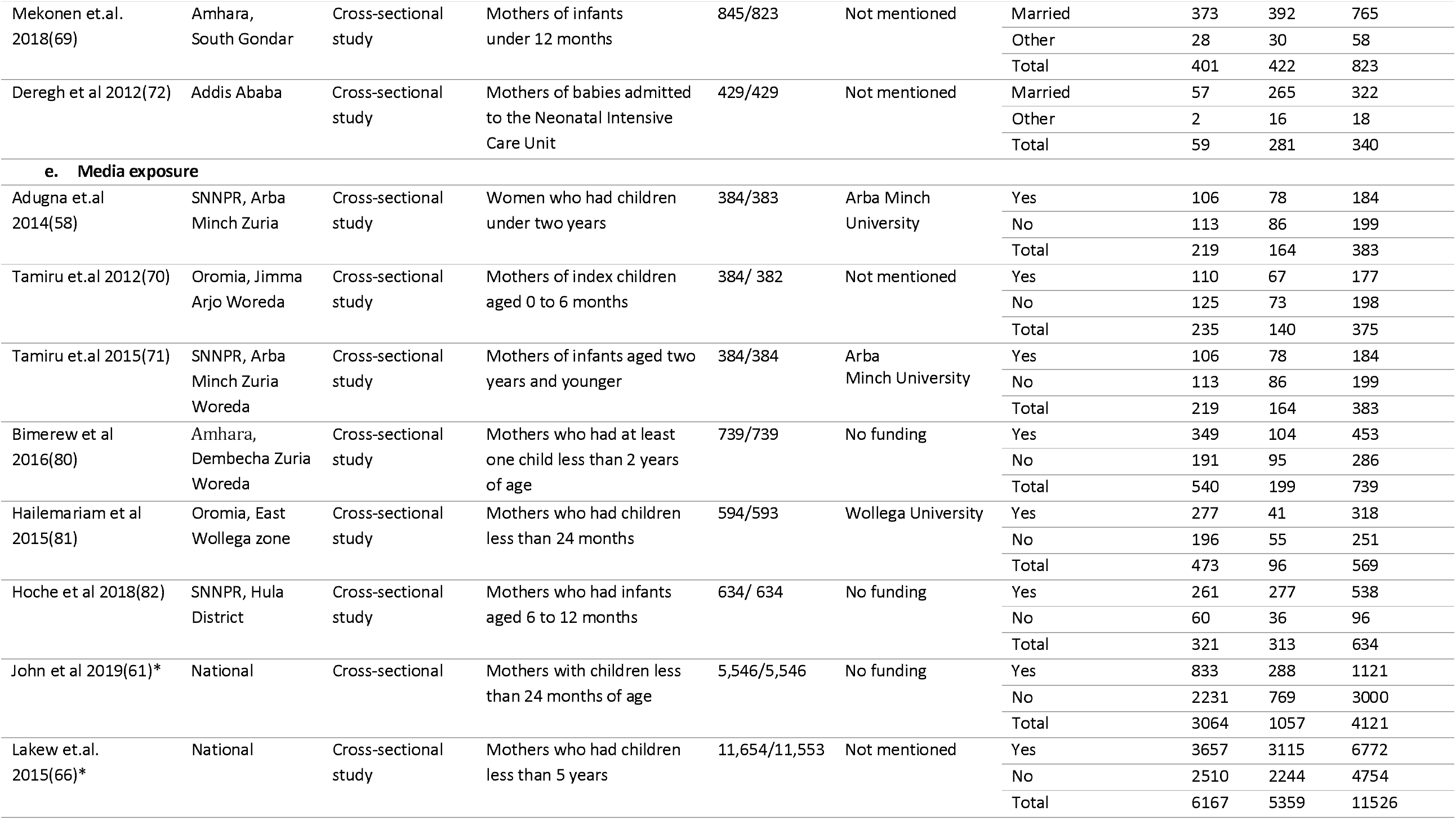
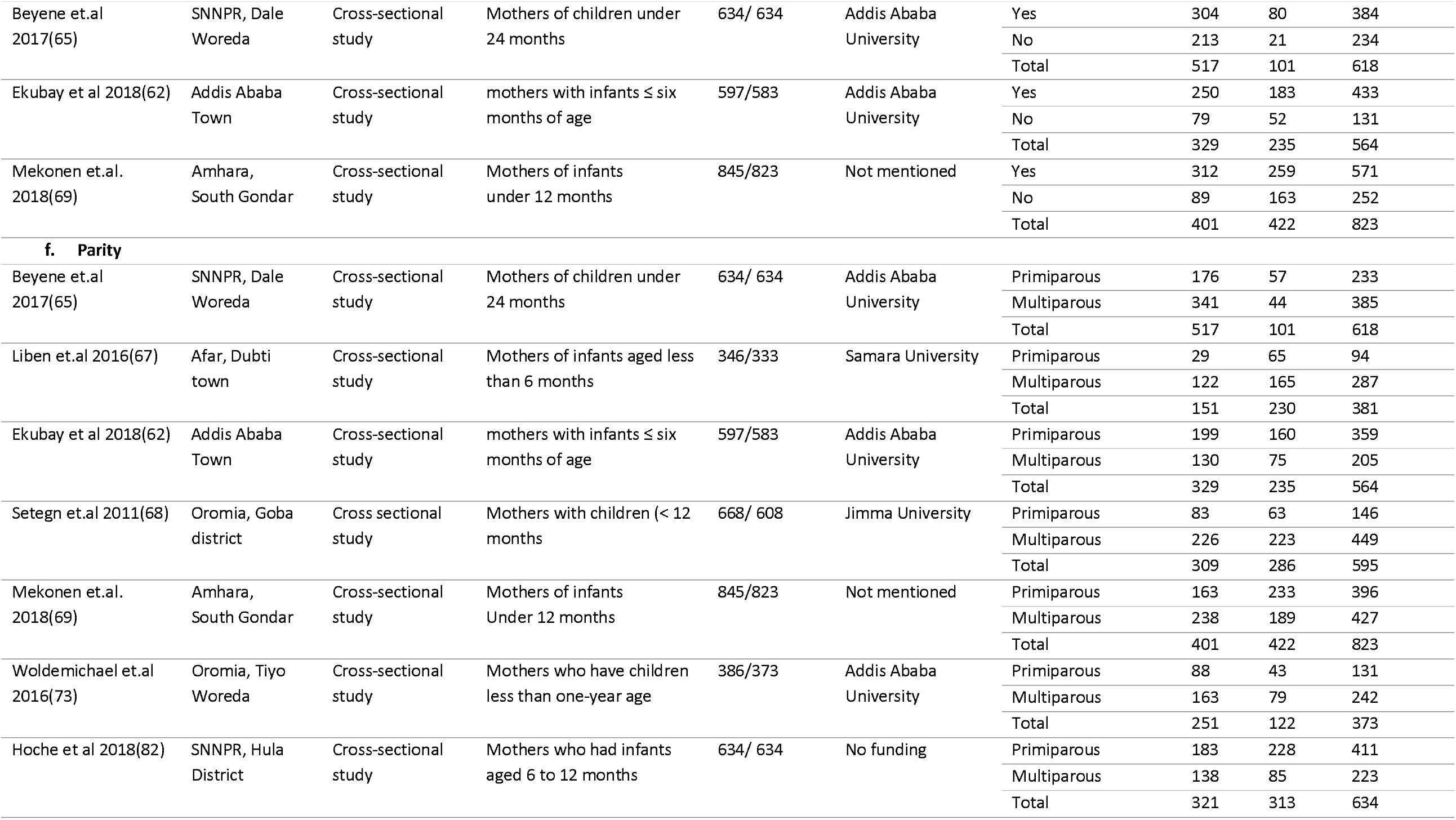
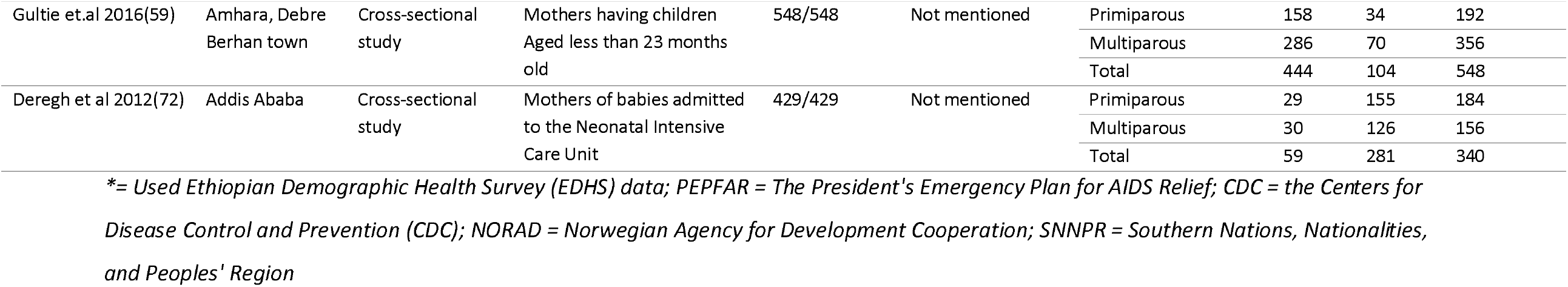
Detailed characteristics of studies that reported each variable of interest.

**Figure 1:**
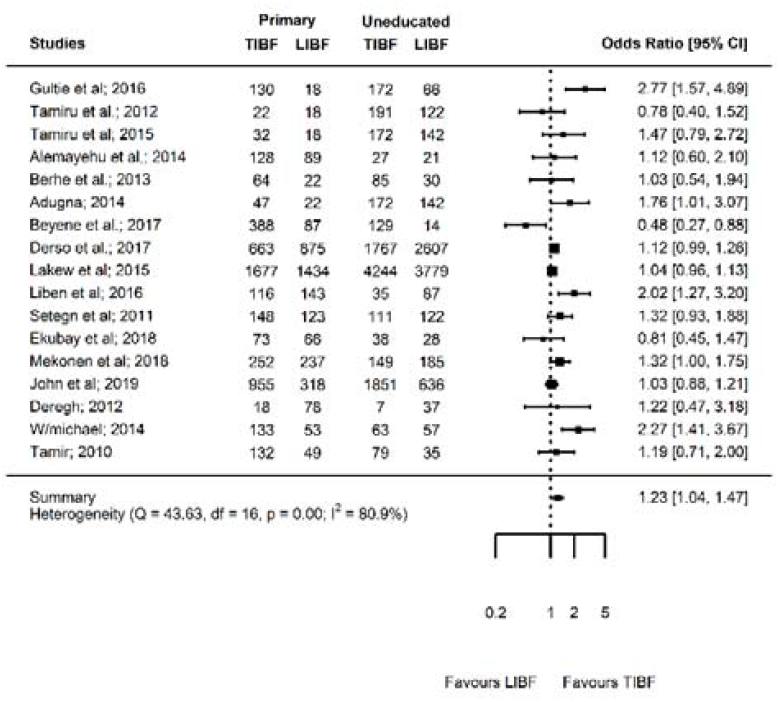
Forest plot showing the results of 17 studies examining the association between maternal educational status (primary education versus uneducated) and timely initiation of breast feeding.

### Predicting factors

#### Maternal educational status

Seventeen studies(58-74) involving 31,066 mothers reported the association between maternal educational status and TIBF (Table 1a). Of which, four studies were conducted in Amhara region, three in Oromia region, three studies in Southern Nations, Nationalities and Peoples Region (SNNPR), and five studies in other regions (Addis Ababa, Afar, and Tigray). Two studies used nationally representative data. The odds of TIBF among mothers who attained primary education was 23% significantly higher than uneducated mothers (OR = 1.23; p = 0.02; 95% CI = 1.04 - 1.47; I^2^ = 80.89%) (Figure 1). The result of meta-regression analysis showed that region, residence, sample size and publication year were not the source of between-study heterogeneity. The odds of TIBF among mothers with secondary education or above was 82% significantly higher than uneducated mothers (OR = 1.82; p < 0.001; 95% CI = 1.35 - 2.45; I^2^ = 84.96%) (Supplementary Figure S1). The meta-regression analysis showed that 78.68% of the between-study heterogeneity explained by variation in region and publication year. Region had significant moderation effect (QM= 22.72, df = 6, p < 0.01). Additionally, the odds of TIBF among mothers with secondary education or above was 43% significantly higher than mothers who attained primary education (OR = 1.43; p = 0.002; 95% CI = 1.15 - 1.79, I^2^ = 73.03%) (Supplementary Figure S2). In all three comparisons, there was no significant publication bias (Supplementary Figures S3, S4, S5) and the evidence on the effect of maternal educational status on TIBF did not markedly change over time (Supplementary Figures S6, S7, S8). Overall, we observed a direct dose-response relationship between maternal educational status and TIBF.

#### Paternal educational status

Four studies(61, 71, 73, 75) with 11,491 mothers reported the association between paternal educational status and TIBF (Table 1b). These studies were conducted in Amhara, Oromia, and SNNPR. One study used nationally representative data. The odds of TIBF in mothers who had a spouse with primary education was 17% higher than mothers who had an uneducated spouse, although the difference was not statistically significant (OR = 1.17; p = 0.56; 95% CI = 0.69 - 2.01, I^2^ = 89%) (Supplementary Figure S9). The meta-regression analysis revealed that 12.68% of between-study heterogeneity due to the difference in residence and publication year of studies, however, moderation effect was not observed. Mothers who had a spouse with secondary education or above were 2.72 times significantly higher chance of TIBF compared with mothers who had an uneducated spouse (OR = 2.72; p = 0.001, 95%CI = 1.49 - 4.97 I^2^ = 62.50%) (Figure 2). This result was obtained after removing one outlier study(61) using Jackknife analysis. Moreover, the odds of TIBF in mothers who had a spouse with secondary education or above was 66% higher than mothers who had had a spouse with primary education although this difference was not statistically significant (OR = 1.66; p = 0.21; 95% CI = 0.75 - 3.66, I^2^ = 90.44%) (Supplementary Figure S10). The meta-regression analysis revealed that 12.68% of between-study heterogeneity can be explained by the difference in the residence but had no moderation effect. In all comparisons, there was no significant publication bias (Supplementary Figures S11, S12, S13). The evidence on the effect of paternal educational status on TIBF was mixed with dramatic decrement (Supplementary Figures S14 and S15) and stability (Supplementary Figure S16) over time. Similar to maternal educational status, there was a clear indication of the dose-response relationship between paternal educational status and TIBF, although the association was not consistent at different grade level.

**Figure 2:**
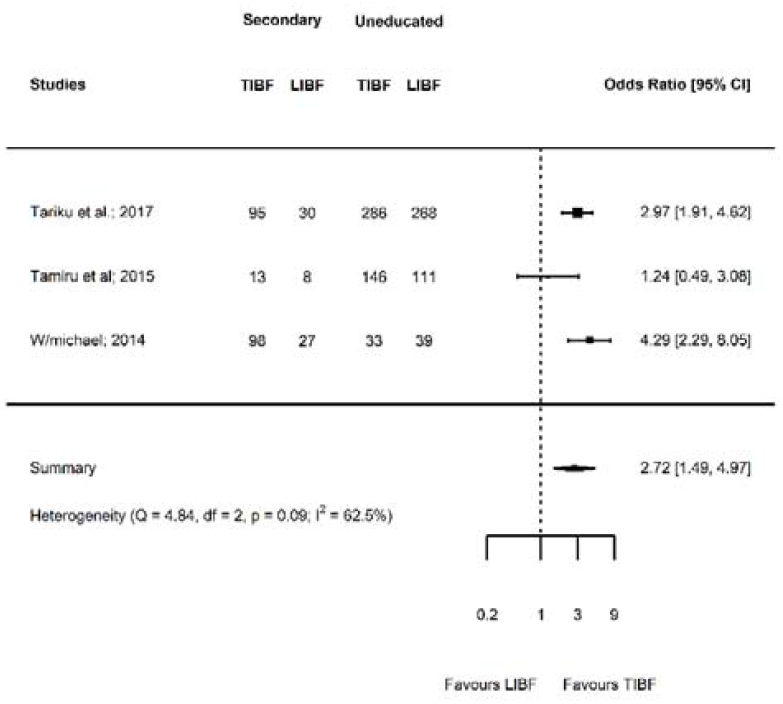
Forest plot showing the results of four studies examining the association between paternal educational status (Secondary or above versus uneducated) and timely initiation of breast feeding.

#### Household income

Eight studies(61, 62, 64, 66, 75-78) including 21,265 mothers reported the association between income and TIBF (Table 1c). Three studies conducted in Amhara region and the remaining three studies were from Addis Ababa, Tigray, and SNNPR. Two studies used nationally representative data. The odds of TIBF among mothers who had medium income was 10% higher than mothers who had low income (OR = 1.10; p = 0.64; 95% CI = 0.75 - 1.60; I^2^ = 92.44%) (Supplementary Figure S17). The meta-regression analysis showed that the heterogeneity fully accounted for variation in region and residence, and these factors had a moderation effect as well (QM= 50.72, df = 5, p < 0.001). The odds of TIBF among mothers who had high income was 28% higher than mothers who had low income (OR = 1.28; p = 0.25; 95% CI = 0.84 - 1.96; I^2^ = 93.87%) (Supplementary Figure S18). The meta-regression analysis showed that 91.50% of the heterogeneity accounted for the variation in the region, residence, and sample size. These factors had also moderation effect (QM= 32.24, df = 6, p < 0.01). The odds of TIBF among mothers who had high income was 16% significantly higher than mothers who had medium income (OR = 1.16; p = 0.002; 95% CI = 1.05 - 1.27; I^2^ = 0.00%) (Figure 3). This was the estimate after excluding one outlier study(61) using Jackknife sensitivity analysis. There was no significant publication bias in all comparisons (Supplementary Figures S19, S20, S21). The evidence on the effect of income on TIBF was slightly increased over time (Supplementary Figures S22, S23, S24). Overall, there was a reverse dose-response relationship between income and TIBF, although the association was not consistently significant.

**Figure 3:**
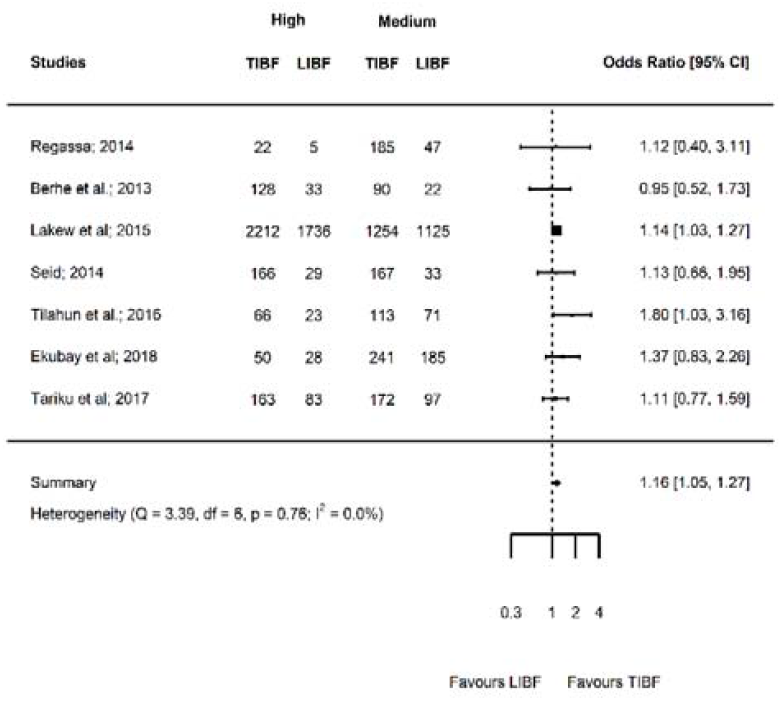
Forest plot showing the results of eight studies examining the association between household income (high versus medium) and timely initiation of breast feeding.

#### Marital status

Twelve studies(60-62, 64, 65, 67-70, 72, 73, 79) involving 17,259 mothers reported the association between marital status and TIBF (Table 1d). Three studies were conducted in Amhara region, three in Oromia region and the rest were conducted in Addis Ababa, Afar, and Tigray while one study was nationally conducted. One outlier study(60) was excluded after Jackknife sensitivity analysis. The odds of TIBF among married mothers was 39% significantly higher than unmarried mothers (OR = 1.39; p = 0.001; 95% CI = 1.14 - 1.69; I^2^ = 9.17%) (Figure 4). There was no significant publication bias (Supplementary Figure S25). The evidence on the effect of marital status on TIBF was slightly increased over time (Supplementary Figure S26).

**Figure 4:**
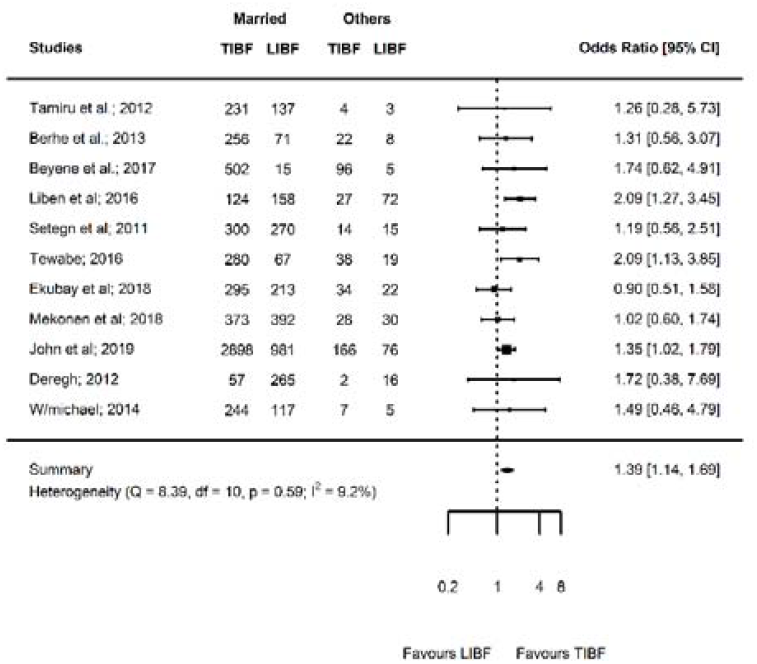
Forest plot showing the results of 11 studies examining the association between marital status (married versus others) and timely initiation of breast feeding.

#### Media exposure

Eleven studies (58, 61, 62, 65, 66, 69-71, 80-82) with 22,315 mothers reported the association between media exposure and TIBF (Table 1e). Four studies were conducted in SNNPR, two studies in Amhara region, two studies in Oromia region and one study in Addis Ababa. Two studies used nationally representative data. The odds of TIBF among mothers who were exposed to media was 5% higher than mothers who were not exposed media, although the difference was not statistically significant (OR = 1.05; p = 0.74; 95% CI = 0.79 - 1.40; I^2^ = 92.15%) (Figure 5). There was no significant publication bias (Supplementary Figure S27). The evidence on the effect of media exposure on TIBF over time was stable (Supplementary Figure S28). The meta-regression analysis result showed that 62.61% of the between-study heterogeneity accounted for the variation in region and publication year, however, none of these factors had moderation effect (QM= 16.44, df = 5, p = 0.05).

**Figure 5:**
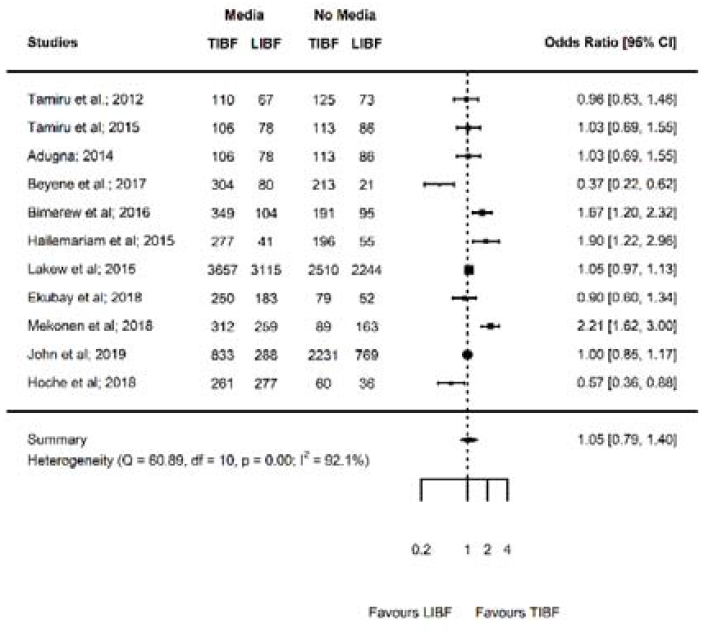
Forest plot showing the results of 11 studies examining the association between media exposure (yes versus no) and timely initiation of breast feeding.

#### Parity

Nine studies(59, 62, 65, 67-69, 72, 73, 82) with 4,993 mothers reported the association between parity and TIBF (Table 1f). Of these, two studies were conducted in Addis Ababa, two in Amhara, two in Oromia, two in SNNPR and one in Afar region. The odds of TIBF among multiparous mothers was 39% significantly higher than primiparous mothers (OR = 1.39; p = 0.01; 95% CI = 1.07 - 1.81; I^2^ = 74.43%) (Figure 6). There was no significant publication bias (Supplementary Figure S29). The evidence on the effect of parity on TIBF was steadily increased over time (Supplementary Figure S30).

**Figure 5:**
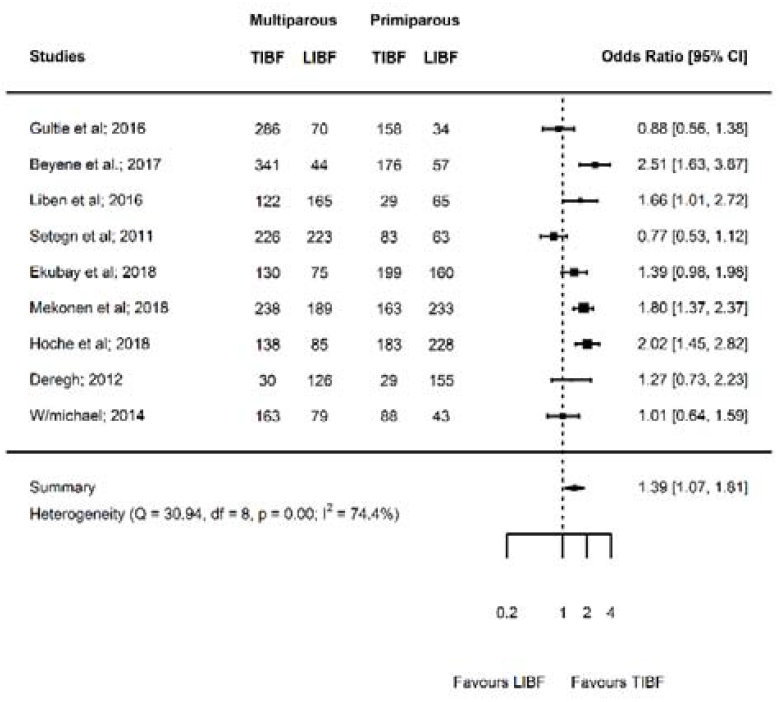
Forest plot showing the results of nine studies examining the association between parity (multiparous versus primiparous) and timely initiation of breast feeding.

## Discussion

This meta-analysis unraveled the association between timely initiation of breast feeding (TIBF) and maternal educational status, paternal educational status, household income, marital status, media exposure, and parity. The key findings were TIBF significantly associated with maternal educational status, paternal educational status, household income, marital status, and parity. We further observed a clear dose-response relationship of TIBF with educational status and household income.

In this study, the odds of TIBF among mothers with primary education was significantly higher than uneducated mothers. The association was even stronger when we compare mothers with secondary education or above with uneducated mothers and mothers who attained primary education, which clearly shows a direct dose-relationship. We observed a similar association between paternal educational status and TIBF. This finding was in line with our hypothesis and studies in India, Nepal, and Timor-Leste.(83, 84) It is likely that education plays an important role in changing the beliefs and attitudes of mothers toward breast feeding, increasing confidence, increase spousal support, maximize ANC follow-up, increase maternal decision making and a higher likelihood of delivering at health institution. In addition, highly educated mothers and fathers could have high expectation, pose questions and request to be properly counselled. On the other hand, other studies(31, 32, 85) reported the absence of a significant association between education status and TIBF. Another study(86) revealed that high educational status found to be associated with delayed initiation of breast feeding. This inconsistency might be attributed to the difference in the study population, sample size and socioeconomic status. The pooled results of our meta-analysis and others (India, Nepal, and Timor-Leste) are based on nationally available studies or data.(83, 84) In addition, delayed initiation of breast feeding in highly educated mothers may be due to the use of private maternities and the larger rate of caesarean section.(87)

In agreement with previous studies(31, 87), we also showed mothers who had high income are likely to pratice TIBF compared with mothers with low income. This might be due to wealthier mothers are more likely to follow the recommended level of antenatal care visits, deliver at health institution and receive adequate postnatal care which can help them to timely initiate breast feeding, whereas poor mothers are deprived of those economic advantages.(88-90) In our previous meta-analyses(30, 40, 41), we affirmed that antenatal care, health institution delivery, and postnatal care significantly associated with optimal breast feeding. In contrast, evidence from Timor-Leste demographic and health survey showed that income was not associated with TIBF.(85)

Moreover, our meta-analysis also showed that currently married mothers timely initiate breast feeding compared with currently unmarried (i.e. single, widowed, divorced) mothers. This finding was supported by previous longitudinal family-based study.(91) This may result from several reasons. Married mothers could have higher incomes and educational levels(92), as we showed socioeconomic advantage increased likelihood of TIBF. In addition, previous study has shown that married mothers are more satisfied, committed to spousal relationships, get shared spousal support and reported less conflict, and therefore, married mothers can have high emotional responsibility to keep the health of the newborn and more likely to engage in positive parenting behaviours.(93) In contrast, a systematic review by Esteves and colleagues reported the absence of association between marital status and initiation of breast feeding in 10 out of 12 studies.(87) This discrepancy might be due to difference in cultural significance of marriage, as a result, the rate of marriage is low or high.

In this study, we further found a significant positive association between multiparity and TIBF. This finding supported our hypothesis and is similar to the evidence in Middle East countries.(32) This may be due to the knowledge advantage that experience give to multiparous women compared with the inexperienced primiparous women. There is evidence that multiparity is associated with better breastfeeding related knowledge.(94) In addition, the increased knowledge and experience may change maternal attitude and behaviour towards breast feeding. On the contrary, a systematic review reported the absence of association between parity and initiation of breast feeding.(87)

In this meta-analysis, we depicted the relevant effect of maternal and paternal education, income, marital status, and parity ob breast feeding initiation. Therefore, health professionals should give special emphasis on uneducated or less educated parents with alternative supportive and educational interventions, such as prenatal education, counseling, and peer education.(83) School education is also required for young girls to increase breast feeding awareness and to prepare for motherhood.(83) Maternal education plays an important role in infant feeding behavioral change and to maintain the mother and newborn health. Even though there has been huge improvements in womens education in Ethiopia, it has long been neglected and remains much lower than men. In addition, interventions targeting fathers at antenatal and postnatal periods may increase breast feeding practices, and should be incorporated in breast feeding programs.(95) Improving the economic power of women would also increase TIBF practice. Furthermore, health professionals should give attention to single and primiparous mothers before, during and after birth. The training of health community agents and structural changes in health services might be relevant to increase TIBF practice.(87)

This meta-analysis showed the strength of association and provided national evidence as previous national, regional, or international studies lacked to report the pooled effect of various relevant factors affecting TIBF. Thus, future researchers in low- and middle-income countries can focus on meta-analysis instead of narrative review. To minimize the possibility of missing relevant studies, we have used a combination of electronic databases search and manual search of cross-references, grey literature, and table of contents of relevant journals. Furthermore, this meta-analysis was conducted based on a published protocol to minimize methodological biases.

We admitted that this meta-analysis has also limitations, which should be taken in to account during the translation of results. All the studies included in this meta-analysis were cross-sectional study, which hinders inference on a cause-effect relationship. The risk of measurement error and recall bias should also be acknowledged. Interestingly, almost all included studies were conducted in mothers with a newborn less than 23 months. In relation to this, the maternal recall is found to be a valid and reliable estimate of breast feeding initiation and duration when the data is collected within three years of breast feeding history.(96) Social desirability bias could also be evident given that self-reported breast feeding experience, educational status and income was used. In addition, the household income classifications are not standardized and did not account for indices such as inflation, which changes over time. Another problem was the confounding effect that can not be excluded given that the reported effect size in this meta-analysis was not adjusted to covariates, such as breast feeding counseling during ANC and PNC follow-up. Studies were lacking in some regions for some variables, which may limit generalizability of our findings. Despite this fact, at least one study used nationally representative data per each variable. In addition, only four studies investigated the association between paternal educational status and TIBF, which may limits the statistical power of our meta-analysis. This meta-analysis only covers studies in Ethiopia; therefore, a comparative meta-analysis from other LMCs and developed countries is required. Moreover, statistical heterogeneity was observed in some of the analysis. Even though residual heterogeneity is still evident, we managed this problem by conducting meta-regression analysis and by removing outlier studies using the Jackknife method. In addition, clinical heterogeneity was minimized by including only studies conducted on healthy mothers and newborns. Methodological heterogeneity was minimized by including studies that reported TIBF based on WHO definition, selected study subjects using a random sampling method, and similar study design and data collection methods. The dose-response relationship of multiparity and TIBF was not investigated due to the huge difference between included studies in the categorization of multiparity. A further limitation of this study was maternal and paternal education represents the formal education gained through schooling and it may not reflect the health literacy of the mothers and fathers. Finally, the results cannot be extrapolated to mothers and newborns with HIV/AIDS or other medical illness.

Measuring breast feeding is challenging, and the use of standardized questions may be interpreted differently according to sociocultural contexts and differing probing techiniques by interviewers.(97) In developing countries, breast feeding coverage is relatively good but we noted studies focused only on factors affecting breast feeding. To brideg this gap, future studies in Ethiopia should focus on investigating the effect of breast feeding on maternal and newborn health outcomes, such as cardiometabolic diseases, neuropsychological diseases, and psychiatric disorders. All included studies in the current review performed the traditional bivariate and multiple logistic regression. As a result, studies on associated factors should deeply consider the interconnection between predictors and apply detailed statistical analysis methods, such as mediation and moderation analysis. For example, it is suggested that geographical, socioeconomic, individual, and health-specific levels factors lower TIBF practice by influencing prelacteal feeding and discarding colostrum, accessibility of information and media, perception, support and milk sufficiency, and involvement of mothers in decision making.(34) Moreover, future research should examine the possible causations between predicting factors and TIBF practices in Ethiopia by implementing longitudinal research designs and large cohort-based studies, which can therefore, produce more accurate and insightful results. Overall, we observed a substantial inconsistency regarding predicting factors across nations; therefore, context-specific meta-analysis is required to strengthen inferences.

To concluded, proximal and distal factors significantly predicting TIBF practice in Ethiopia, which needs integrated intervention by health professionals and healthcare policymakers. Health education, counselling and peer education targeting parents at antenatal and postnatal periods are needed. It is also relevant to improve the economic power of women and promote gender equality.

## Data Availability

All data generated or analyzed in this study are included in the article and its supplementary files.

## Acknowledgment

We would like to Sjoukje van der Werf (librarian at the University Medical Center Groningen, the Netherlands) for her expert advice on search strategies and Balewgizie Sileshi (University of Groningen, the Netherlands) for his help to screen titles and abstracts.

## Data sharing statement

All data generated or analysed in this study are included in the article and its supplementary files.

## Contributors

TD conceived and designed the study, analyzed the data and interpreted the results. TD and SM extracted the data and wrote the first draft of the manuscript. All authors involved in writing, revising and providing intellectual comments, and approved the final manuscript.

